# Improvement of speed-accuracy tradeoff during practice of a point to point task in children with secondary dystonia

**DOI:** 10.1101/2023.05.11.23289830

**Authors:** Maral Kasiri, Emilia Biffi, Emilia Ambrosini, Alessandra Pedrocchi, Terence D. Sanger

## Abstract

The tradeoff between speed and accuracy is a well-known constraint for human movement, but previous work has shown that this tradeoff can be modified by practice, and the quantitative relationship between speed and accuracy may be an indicator of skill in some tasks. We have previously shown that children with dystonia are able to adapt their movement strategy in a ballistic throwing game to compensate for increased variability of movement. Here we test whether children with dystonia can adapt and improve skill learnt on a trajectory task. We use a novel task in which children move a spoon with a marble between two targets. Difficulty is modified by changing the depth of the spoon. Our results show that both healthy children and children with secondary dystonia move more slowly with the more difficult spoons, and both groups improve the relationship between speed and spoon difficulty following one week of practice. By tracking the marble position in the spoon, we show that children with dystonia use a larger fraction of the available variability, whereas healthy children adopt a much safer strategy and remain farther from the margins, as well as learning to adopt and have more control over the marble’s utilized area by practice. Together, our results show that both healthy children and children with dystonia choose trajectories that compensate for risk and inherent variability, and that the increased variability in dystonia can be modified with continued practice.

## INTRODUCTION

The speed-accuracy tradeoff known as Fitts’ law is ubiquitous in human movement^1^. This law is typically formulated as a relationship between the speed of movement and the endpoint accuracy following a rapid or ballistic movement to target^1,2^. While many possible explanations have been proposed, one of the more enduring possibilities is that the tradeoff represents compensation for activity-dependent noise (signal-dependent noise), so that moving more slowly may reduce noise and permit increased accuracy^2–7^.

More recently, it has been shown that Fitts’ law also applies in tasks where the accuracy of the trajectory matters rather than only the accuracy at the final target^6^. Moreover, the quantitative relationship between speed and accuracy for any individual may be modifiable by practice, suggesting that this relationship may be related to the skill of performance in trajectory-following tasks. A trajectory-following task involves a pre-specified movement and may not fully capture the subject’s ability to plan a trajectory^2,6^. Therefore, we have developed a novel task in which subjects must transport a marble in a spoon from one target to another^6^. By varying the depth of the spoon, the task can be made difficult, and subjects are free to choose not only the speed of movement but the complete velocity profile of the trajectory in order to reach the target as rapidly as possible. The size of the target is not varied, so the choice of trajectory is entirely determined by the spoon difficulty. We have previously shown that this task exhibits a robust speed-accuracy tradeoff^6^, that is the participants slow down performing the more difficult tasks to not drop the marble.

Previous work has shown that the speed-accuracy tradeoff for a trajectory-following task can be modified with practice^6,8,9^. Here we test whether this tradeoff can also be modified by practice for the marble-spoon task by analyzing the marble trajectory in the spoon area. We test this in both healthy children and in children with secondary dystonia^10,11^. Dystonia is a disorder characterized by increased variability of movement and increased signal-dependent noise^5,6,10–12^. Children with dystonia are aware of their increased noise and compensate appropriately in a ballistic target task^3–6,13^. Here we test how children in these two groups with different levels of signal dependent noise adapt to the trajectory-following task by investigating if they adapt their strategy in order to lower the risk of movement^14,15^. We also compare how performance in the two groups is affected by learning during practice over a period of five days.

## MATERIALS AND METHODS

### Subjects

A total of twenty-one children and adolescents (ages15.5 ±3.4 years) performed the study with their preferred (dominant or less dystonic) arm. Eight (5 females and 3 males) were healthy subjects, while thirteen (1 female and 12 males) were diagnosed with secondary (acquired) dystonia. Subjects were diagnosed by a pediatric movement disorder specialist using standard criteria in Children’s hospital, Los Angeles (CHLA) and in IRCCS Medea, Italy^16^. All patients provided signed informed consent for Health Insurance Portability and Accountability Act (HIPAA) authorization if they were recruited in CHLA for the research use of protected health information. Parents of participants recruited at IRCCS Medea signed a written informed consent. The protocol of the study was approved by the IRB and the Ethical Committees of the Scientific Institute E. Medea (reference number: 054/14-CE; Date: 01-04-2015) and CHLA. The experiments took place in two locations, University of Southern California (USC), and IRCCS Medea. Details of the participants with dystonia are provided in Table 1.

**Table 1.**
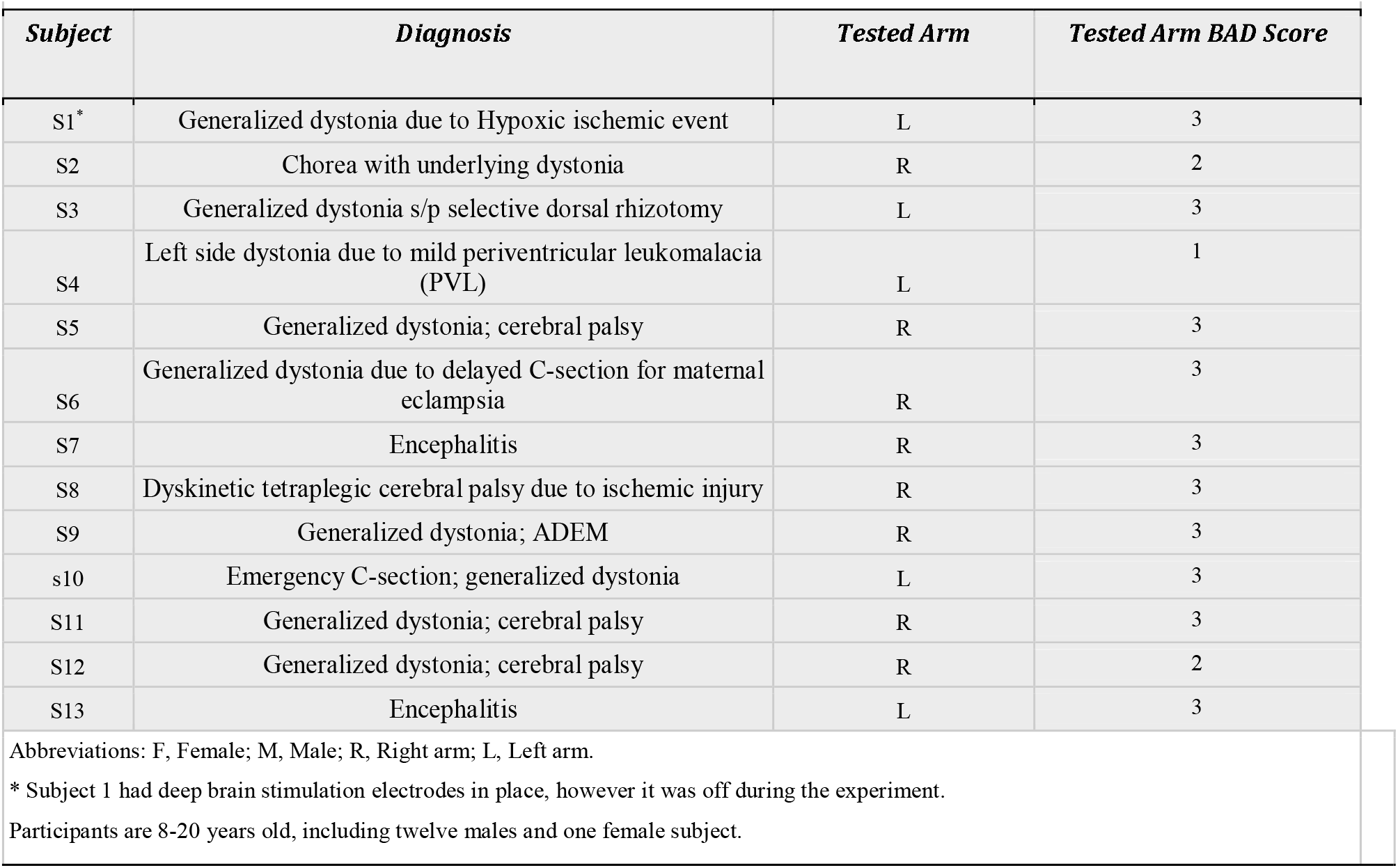
Patients’ demographics.

### Testing

Participants seated upright on an adjustable chair or their own wheelchair. A board with two targets was placed on a desk in front of them. Each target was bounded by two plastic blocks and the distance between the center lines of these targets was 20 cm along the vertical axis^6^ (Figure 1a). The board’s position was adjusted in a way that the subjects had to extend their arm fully to reach the more distant target (Figure 1b). To track the spoon and upper body movement two different systems were used.

**Figure 1.**
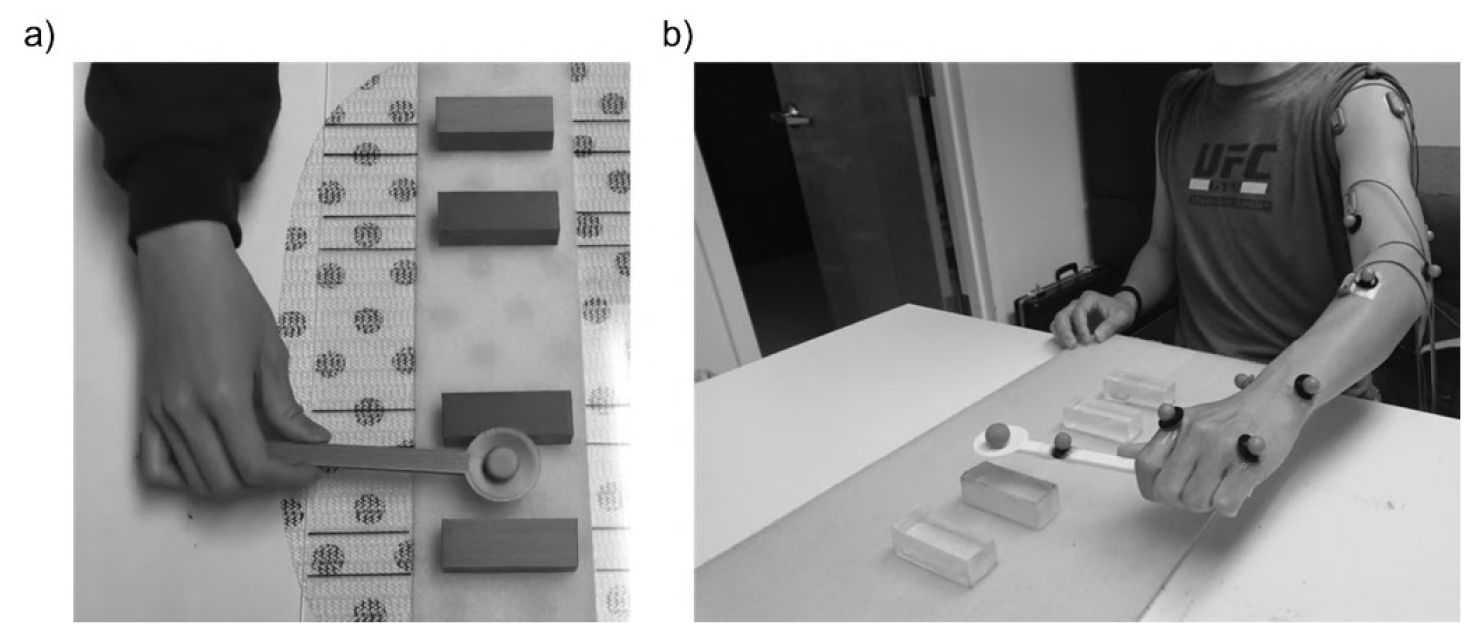
Experiment setup: a) Two plastic blocks are attached to the board for each target, the distance between center lines of targets is 20 cm along the vertical axis. b) 12 passive reflective markers were placed on the upper limb as shown with a additional marker attached to the spoon and another one as the marble carried by the spoon. The participants were asked t extend their arm and the board position was adjusted in a way that the spoon was aligned with the distant target.

At USC four Vicon Nexus 1.8.5 motion capture cameras (© Vicon Motion Systems Ltd, UK) were used to track the spoon and upper body movements. Cameras were placed in four spots in front of the subject and were calibrated using a calibration wand to make sure they can detect all the markers. At Medea, eight optoelectronic cameras by BTS Bioengineering were placed around the subject, calibrated with a calibration wand, to track the spoon and upper body movements. Twelve passive reflective markers were attached to the upper extremity joints and body as shown in Figure 1b. Two additional untethered spherical markers were used, one attached to the spoon and the other as the marble carried by the spoon (total of 14 markers). The subjects were instructed to hold the reflective marble in the spoon and move the spoon back and forth between the two targets as fast as they could without dropping the marble. The upper extremity kinematic data along with the spoon and marble trajectory were recorded at 100 Hz in USC or 60 Hz in Medea to evaluate the participants’ motor performance.

Nine different circular spoons of varying depth were designed in SolidWorks^®^ 2016 (Dassault Systems SOLIDWORKS Corporation) and were 3D printed. The detail of their size is provided in Table 2 and the index of difficulty^1^ for each spoon was calculated based on the spoon dimensions. For each spoon, we calculate the “index of difficulty (ID)” as the ratio of the spoon inner diameter to its depth, noting that the marble size and the spoon diameter is constant. Therefore, the ID is only dependent to the depth of each spoon. Note that unlike in standard Fitts’ law formulations^1^, the ID used here reflects a property of the spoon, not a property of the target (e.g. target size or distance)^6^. Furthermore, while increasing ID reflects the task difficulty at a given speed, there is no simple relationship between ID and performance across all subjects, in part because there is a ceiling effect for the easiest spoons, and some subjects are unable to perform the task at all for the hardest spoons. Therefore, we do not expect a linear relationship between ID and speed of movement.

**Table 2.**
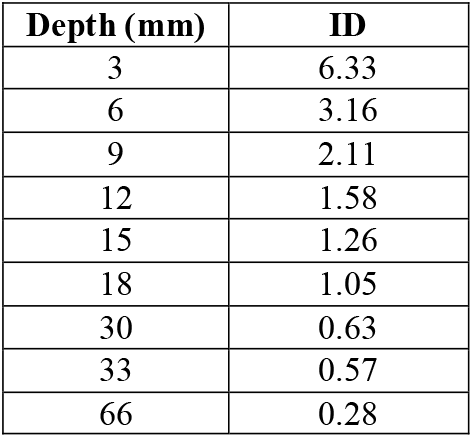
Details of spoon depths and their computed index of difficulty. The inner diameter of each spoon is 19mm. The index of difficulty is then computed as the ratio of diameter to depth of each spoon.

| Depth (mm) | ID |
| --- | --- |
| 3 | 6.33 |
| 6 | 3.16 |
| 9 | 2.11 |
| 12 | 1.58 |
| 15 | 1.26 |
| 18 | 1.05 |
| 30 | 0.63 |
| 33 | 0.57 |
| 66 | 0.28 |

Each subject participated in the experiment for 5 consecutive days. On days one and five, both practice and performance testing occurred in order to determine baseline and improvement with practice. On days 2, 3, and 4, only practice occurred. Prior to initiating the experiment for each subject, performance was tested on a range of different spoon difficulties. Easy, medium, and difficult spoons sizes were chosen for each subject. The difficult spoon was chosen as the largest ID for which the subject could successfully transport the marble dropping it on fewer than 30% of trials. The medium and easy spoons were the next 1 and 2 spoon difficulties below. Testing is performed on all three spoon sizes as well as the spoon without a marble, but training was performed only on the medium spoon size for each subject.

Testing on days 1 and 5 includes 2 trials of 10 repetitions of forward and backward movements with all three spoon sizes. On each trial if they drop the marble more than two times, they had to do the trial again. However, no specific cost was associated with dropping the marble. Training on days 1 through 5 included 5 sets of 10 repetitions of the same task using only the medium spoon size. In this study, we used the kinematic recordings from the testing on days 1 and 5.

### Speed-accuracy trade-off (Fitt’s law)^1^

We first tested the effect of ID on speed and determined how this effect changed following practice. We fitted a regression line to the movement time (MT) and the index of difficulty (ID) and calculated the index of performance (IP) as the inverse of the MT-ID linear equation slope for each subject. The index of performance^1,6^ is used as a measure to evaluate if their performance improved in day five compared to day one. Statistical analyses were done by lme4^17^, emmeans^18^ packages in R-studio (R core team, 2021). A Linear mixed effects model with repeated measures was employed to analyze the effect of practice on the performance.

We assessed the effect of ID, group (i.e., dystonia or healthy), and testing day (i.e., pre or post), and their interactions as independent variables (fixed effects) on MT and speed of movement (dependent variables) by fitting a linear mixed effect model^17^ to the data. In our model, random effects are assumed to be intercepts for subjects and by-subject random slopes for the effect of ID. In the MT model, we assumed uncorrelated slopes and intercepts (equation (1)); however, we assumed correlated slopes and intercepts for the speed of movement linear model due to the singularity of fit (equation (2)). We then performed analysis of variance test (Anova) and pairwise comparison to obtain the significance of each effect.

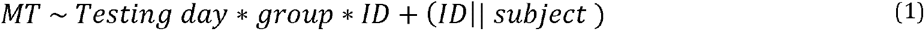

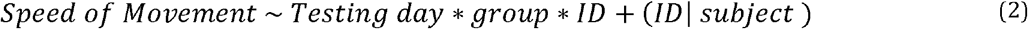

### Marble kinematic analysis

If movement speed is limited by the risk of dropping the marble, then speed can be maximized by allowing the marble to move as much as possible within the spoon. We thus analyzed the marble position within the spoon during movement in order to determine whether maximal areas were used, or whether other considerations such as reduction of risk or physical constraints on movement speed were affecting the performance. The position of the marble inside the spoon lies approximately within an ellipse; thus, an ellipse was fitted to the marble trajectory for each repetition (one forward and one backward movement). This method enabled us to estimate how much of the spoon area is used to achieve the desired accuracy or speed.

The eigenvalues of the movement trajectory (the ellipse diameter along the semi-major and the semi-minor axes) were then computed. We computed a safety margin with the two main eigenvalues, ‘a’ and ‘b’, as a measure to investigate the variability along the first eigenvector of the movement trajectory (e1), and along the second eigenvector of the movement trajectory (e2), respectively. This safety margin is computed based on equations (3) and *(4)* to determine how sensitive each subject is to the risk of movement, before and after practice. Therefore, a smaller safety margin means that the subject is risk-seeking and therefore they use more of the available variability. Similarly, a larger safety margin means that the subject is risk-averse, and they use less of the available variability^15^.

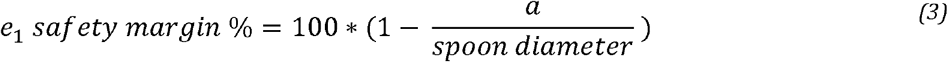

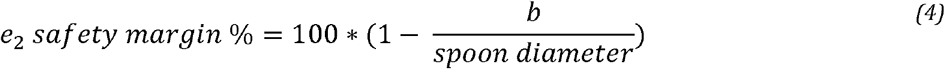

We explored how these ratios change with respect to speed of movement, ID and with practice, for each subject. We employed a repeated measure analysis with linear mixed effect model to derive the statistical results of the change in these ratios with respect to the speed and ID with practice. In the mixed effect model, ID, testing day, speed, and their interactions are the fixed effects. Similar to previous models, random effects are the intercepts for subjects and by-subject random slopes for the effect of index of difficulty (equation (5)). An analysis of variance test (Anova) and pairwise comparison were performed to obtain the significance of each effect.

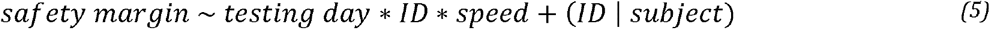

### Smoothness and Coefficient of Variation analysis

In addition to analyzing the marble trajectory and movement time, we assessed the effect of practice on change in smoothness and variability. Dimensionless jerk score is a useful measure to investigate movement smoothness which was computes based on equation _(6)_.

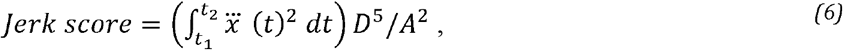

In which *D = t*_*2*_ – *t*_*1*_ is the duration and *A* is the amplitude of movement (distance)^19^. Additionally, the coefficient of variation was derived as the ratio of the speed standard deviation to its mean for each repetition 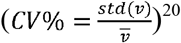.

Similarly, a linear mixed effect model with repeated measures analysis was used to compare the smoothness and the coefficient of variation (CV) with respect to the groups, ID, testing day and their interactions.

## RESULTS

### Speed-accuracy trade-off

We fitted a linear mixed effect model to assess the effect of task difficulty on MT and speed of movement (Figure 2a). The marble trajectory and the fitted ellipse for a patient with dystonia and a healthy subject is shown in Figure 2b. This figure illustrates how they adjust the trajectory with respect to the movement time and index of difficulty.

**Figure 2.**
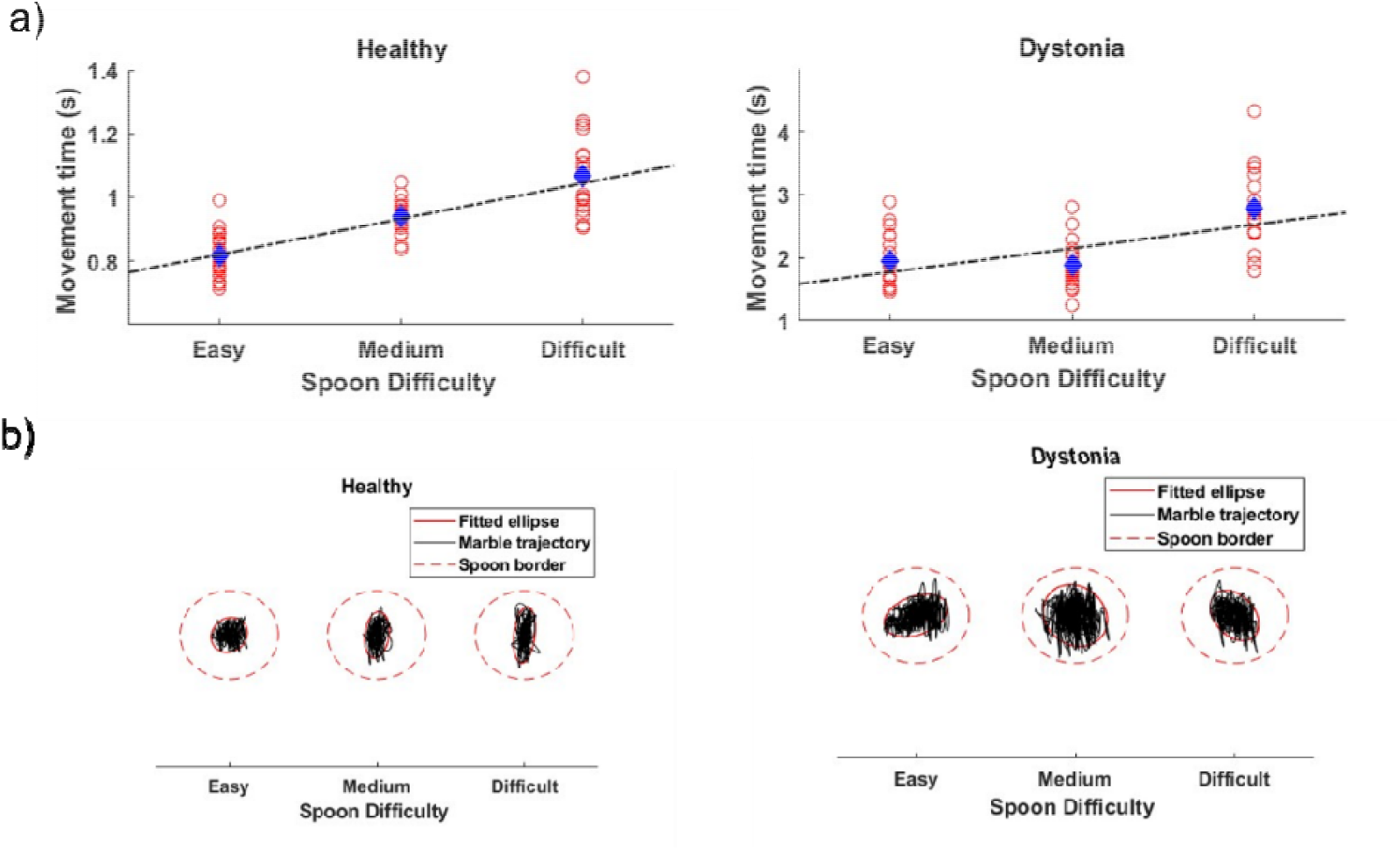
a) The movement time with respect to spoon sizes (easy, medium and difficult) for a subject with dystonia (right) and a healthy subject (left) is shown with red dots for each repetition. The blue dot shows the average movement time for all repetitions in one trial and the dashed black regression line shows the increasing trend of movement time with respect to the spoon difficulty. b) The marble trajectory (black line) within the spoon border (dashed red line) and the fitted ellipse (solid red line) to that trajectory is shown for each spoon difficulty for one healthy subject (left) and one subject with dystonia (right) in one trial.

As illustrated in Figure 3 and consonant with earlier results on this task^6^, the movement speed decreases in all subjects with the increase in task difficulty. Task difficulty is not a limiting factor for the speed of movement as the movement speeds without the marble (ID = 0 in Figure 3) are faster than the other task difficulties. Within group comparison of movement speed showed that the subjects with dystonia (R^2^ _ID_= 0.88, p-value < .001) had a significant increase in their speed of movement in all three spoons with practice (day 1 versus day 5, Figure 4). There was no significant change in the speed of movement of healthy children with the easier spoon, however the model predicted an increase in speed with practice in higher task difficulties (medium and difficult) ([R^2^_spoon_= 0.88, p-value _easy_ = .48, p-value _medium_ = .02, p-value _difficult_ = .04], [R^2^ _ID_= 0.77, p-value <.05]). Figure 4 shows the average speed and the standard deviation in day 1 versus day 5 for the healthy children and those with secondary dystonia in all three task difficulties. Similar results were obtained by performing anova using type II Wald chi-square test for dystonia group (Pr_(>chisq)_ < .001) and for healthy group (Pr_(>chisq)_ < .05), showing the significant effect of index of difficulty and the testing day on the speed of movement.

**Figure 3.**
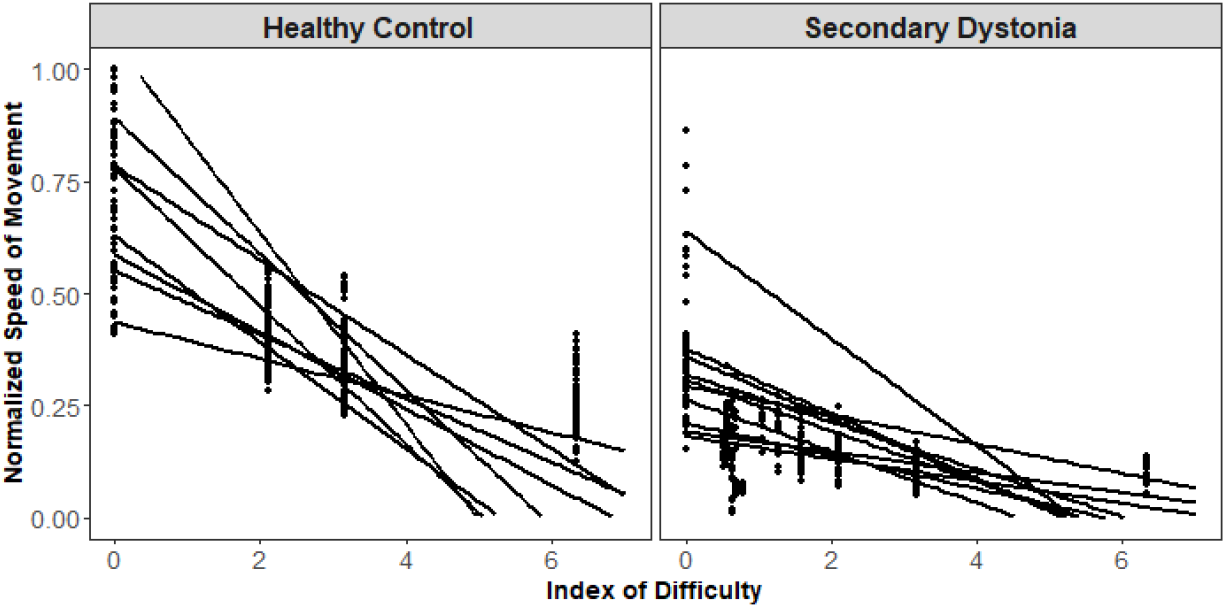
Normalized speed of movement versus the index of difficulty (ID) (higher ID indicates the more difficult task and zero ID indicates the no-marble condition.) for the healthy control and dystonia group is shown in dots. Each dot represents the speed for one repetition of task with the corresponding ID, before the training (day 1). Regression lines for the speed of movement versus the ID for each subject (p-value < 0.01 for all except two children with dystonia) are shown for both groups. The decreasing trend of movement with respect to the ID indicates that all the subjects follow the Fitt’s law and adjust their speed based on the task difficulty.

**Figure 4.**
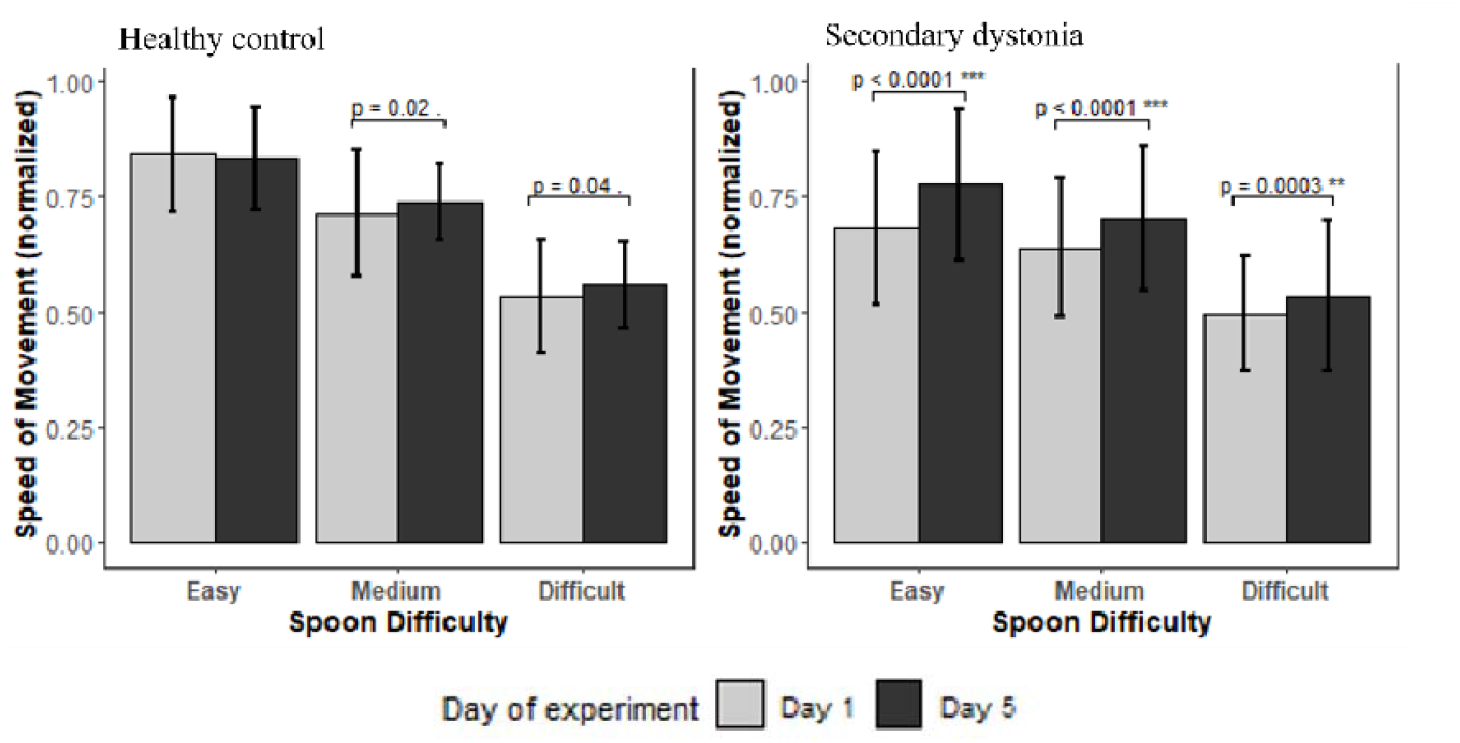
Statistical result of the improvement in the speed of movement in each group with three different task difficulties. The figure depicts the averaged normalized speed of movement and the standard deviation versus the testing day for three spoon sizes in healthy (left) and dystonia (right) group, predicted by the linear mixed effect model.

The regression lines of the movement time versus index of difficulty for day one and day five is shown for in each group in Figure 5 (only three subjects are shown for clarity). All the regression lines except two patients with secondary dystonia had a slope significantly different from zero (P-value < .05), meaning the MT changed significantly with the difficulty (they followed the Fitt’s law of speed accuracy trade-off). The pairwise comparison to estimate the marginal means for the fitted model on the movement time revealed a significant improvement in the index of performance (inverse of the slopes) in healthy group (R^2^_ID_ = 0.73, p-value = 0.001) and the dystonia group (R^2^_ID_ = 0.89, p-value < 0.001) with practice (Figure 6). Analysis of variance (anova) using type II Wald chi-square test performed on the linear models revealed a significant effect (Pr_(>chisq)_ < .0001) of testing day and ID but not their interaction (testing day * ID).

**Figure 5.**
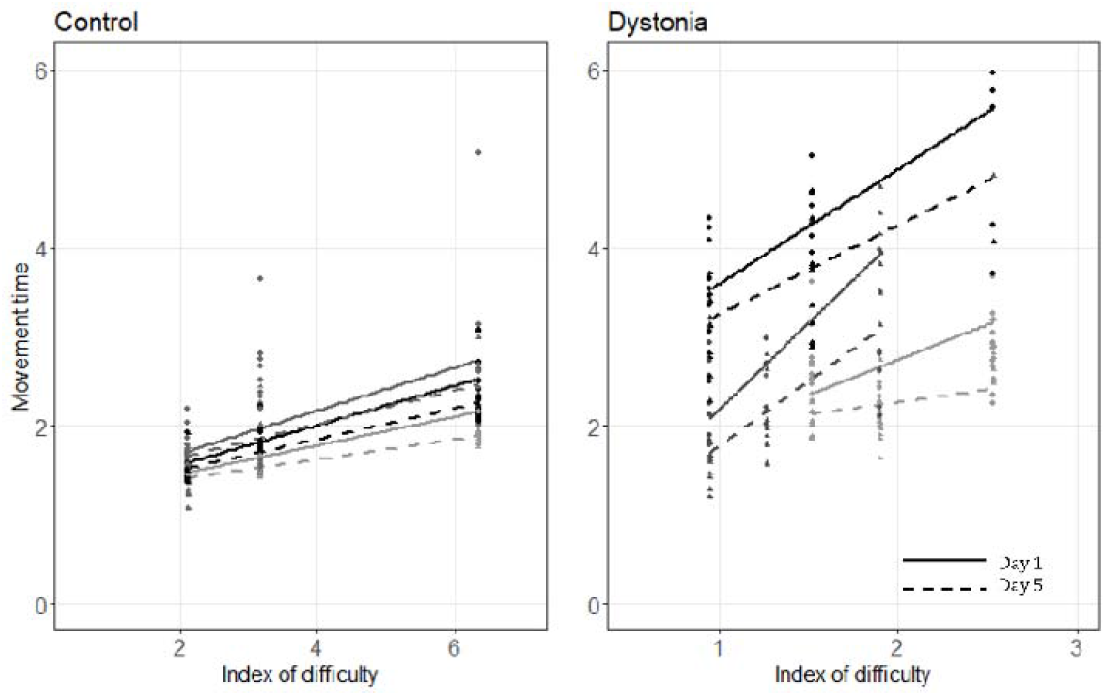
Change in movement time (MT) for three participants in each group (healthy control and dystonia) with respect to ID for day 1 (solid line) and day 5 (dashed line). Please note that the ID axis scale is different for healthy children and children with dystonia due to their different capabilities. The slope of each line indicates the inverse of index of performance for the corresponding subject on either day 1 (solid line) or day 5 (dashed line).

**Figure 6.**
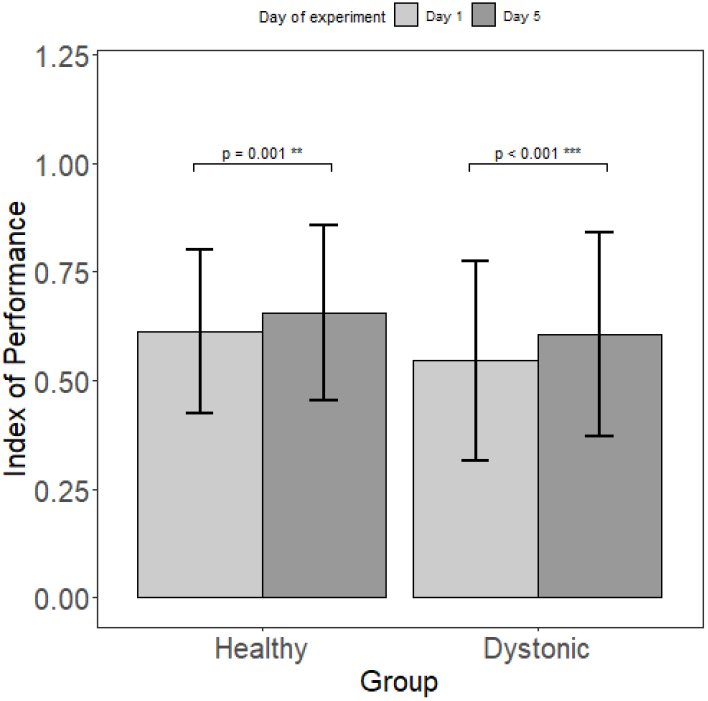
Statistical result of the normalized IP comparison before and after training in each group. Increase in the IP measures the effect of practice on learning.

Based on the protocol, the spoon sizes that each subject practiced with are consistent throughout the whole experiment from day 1 to day 5; therefore, changes in IP are entirely due to changes in the speed of movement. Movement speed improved significantly for each of the spoon sizes individually in the children with dystonia, but the change in speed for the healthy children was only significant when combining performance across multiple spoon sizes.

### Marble Kinematics Analysis

#### The safety margin in the direction of first eigenvector (e_1_ safety margin)

The statistical tests performed on the e_1_ safety margin reveals that there was a significant decrease of e_1_ safety margin with practice in healthy control group. The linear model explains 76% of variance in the data (R^2^= 0.76) and the anova test revealed that all the variables and their interactions except day*movement speed have significant effect on the e_1_ safety margin (Pr_(>chisq)_ < .001).

On the other hand, the effects of these parameters on the e_1_ safety margin were shown to be negligible in children with dystonia. The fitted linear mixed effect model (R^2^= 0.57) and the performed anova test showed that there is no significant change in e_1_ safety margin with practice and with the change of movement speed or ID, in children with dystonia. These results are illustrated in Figure 7 with respect to spoon difficulty for each group. Figure 8 shows the regression lines for three subjects in each group with respect to ID and speed of movement, respectively.

**Figure 7.**
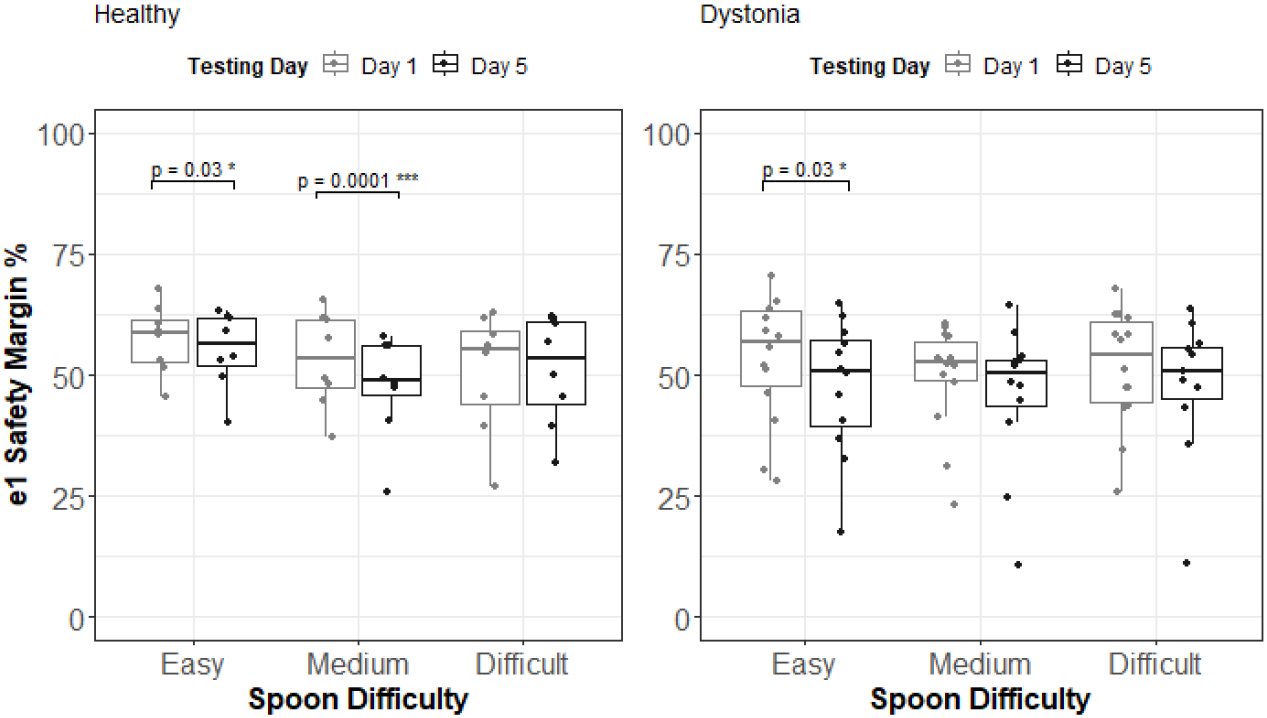
e1 safety margin group analysis for healthy control group and dystonia group. This ratio does not change with respect to index of difficulty; however, pairwise comparison revealed that healthy control group showed a significant decrease in this ratio with practice, performing with easy and medium spoon difficulty; and this ratio only decrease in children with dystonia performing with easy spoon difficulty.

**Figure 8.**
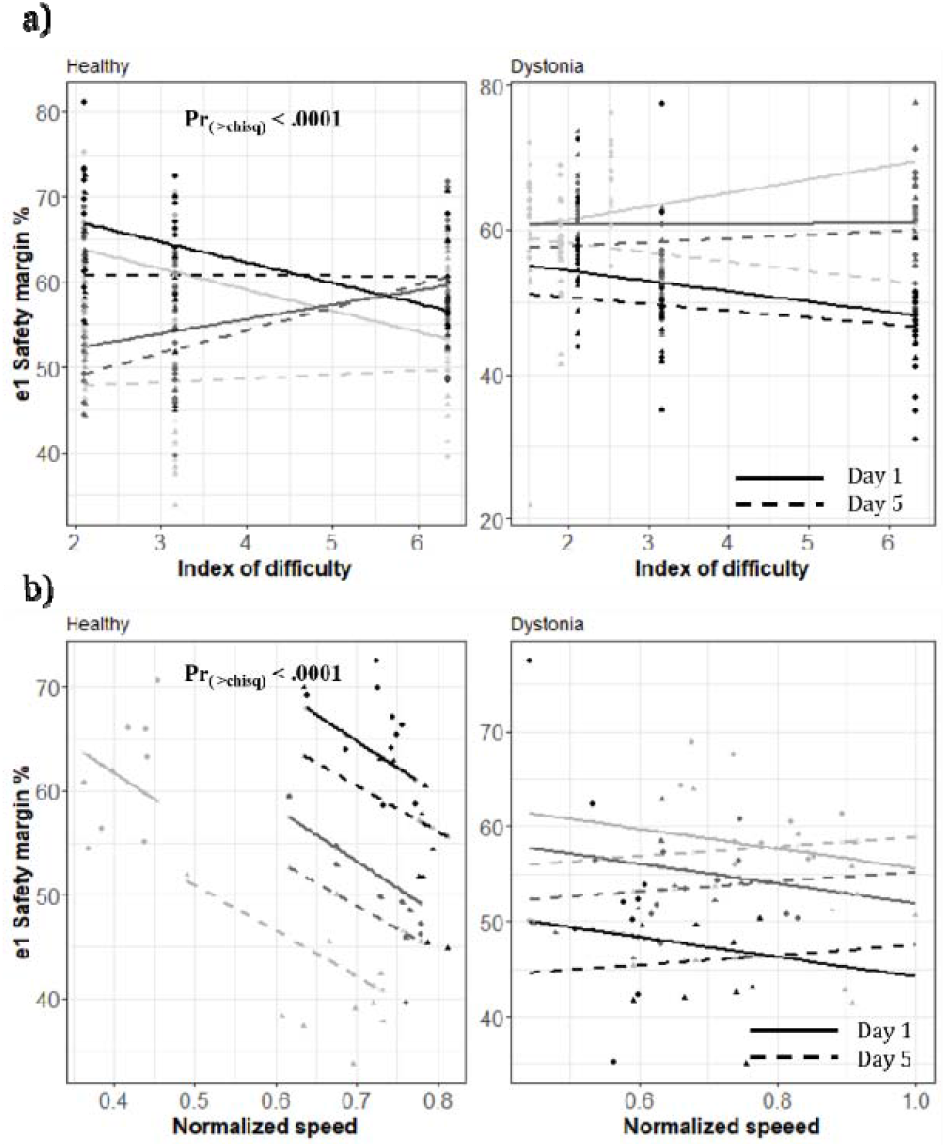
Fitted linear model: a) This figure shows the fitted lines for three subjects (three shades of gray) from ach group with respect to ID. The fitted lines are consistent with earlier data shown in Figure 7; we see a higher decrease in this ratio in smaller indices of difficulty in healthy control group (Pr (>chisq) < .0001). b) This figure shows the same fitted lines for those subjects with respect to speed of movement. It clearly shows that the slopes are significant (Pr (>chisq) < .001) for healthy control group, as well as the extent of drop in this ratio.

#### The safety margin in the direction of second eigenvector (e_2_ safety margin)

We fitted linear mixed effect models and performed pairwise comparison on them for the healthy control group (R^2^ =0.49, p-value = 0.50) and the dystonia group (R^2^ = 0.77, p-value= 0.43). The analysis of variance showed only a significant effect of ID * speed (Pr_(>chisq)_ < .0001) for healthy control group (Figure 9). No other significant effects were observed on e_2_ safety margin as illustrated in Figure 9.

**Figure 9.**
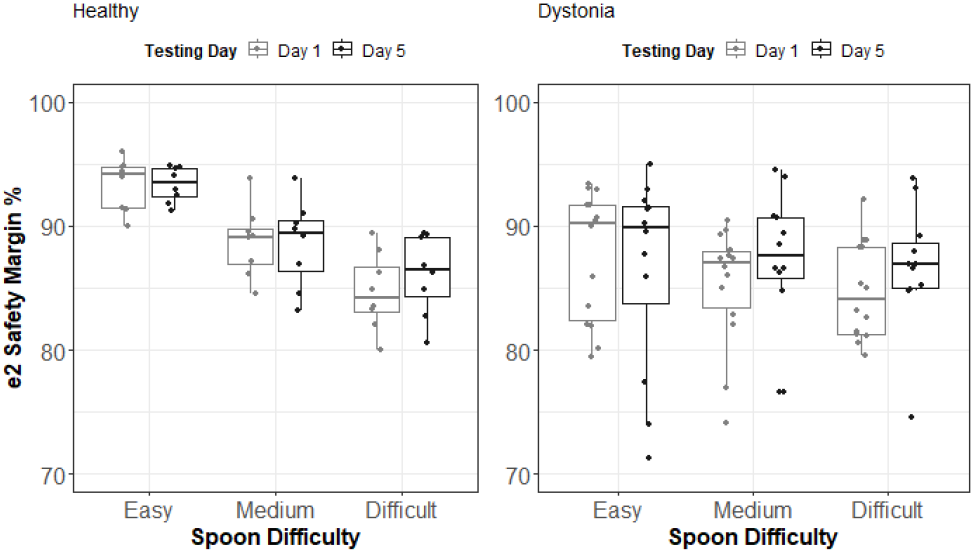
e2 safety margin group analysis for healthy control group and dystonia group. There was no change in this measure associated with practice; however, in healthy control group there is a significant decay of this measure with respect to spoon difficulty, consistent with the results of anova on the effect of ID * speed of movement (Pr(>chisq) < .001); The e2 safety margin decreases as the ID increases in healthy control group.

### Smoothness and Coefficient of Variation Analysis

We performed the same analysis on the jerk scores in both groups. The models explained 79% and 53% or variance of jerk score in healthy control and the dystonia group, respectively. The analysis of variance showed a significant decrease of jerkiness (increased smoothness) with practice in children with dystonia (Pr_(>chisq)_ < .001) for the easy task difficulty (Figure 10a). The result is consistent with the pairwise comparison of means which revealed a significant decrease of jerkiness with practice (p-value = .01) in children with secondary dystonia with the easy task (Day 1 mean = 0.4; Day 5 mean = 0.14).

**Figure 10.**
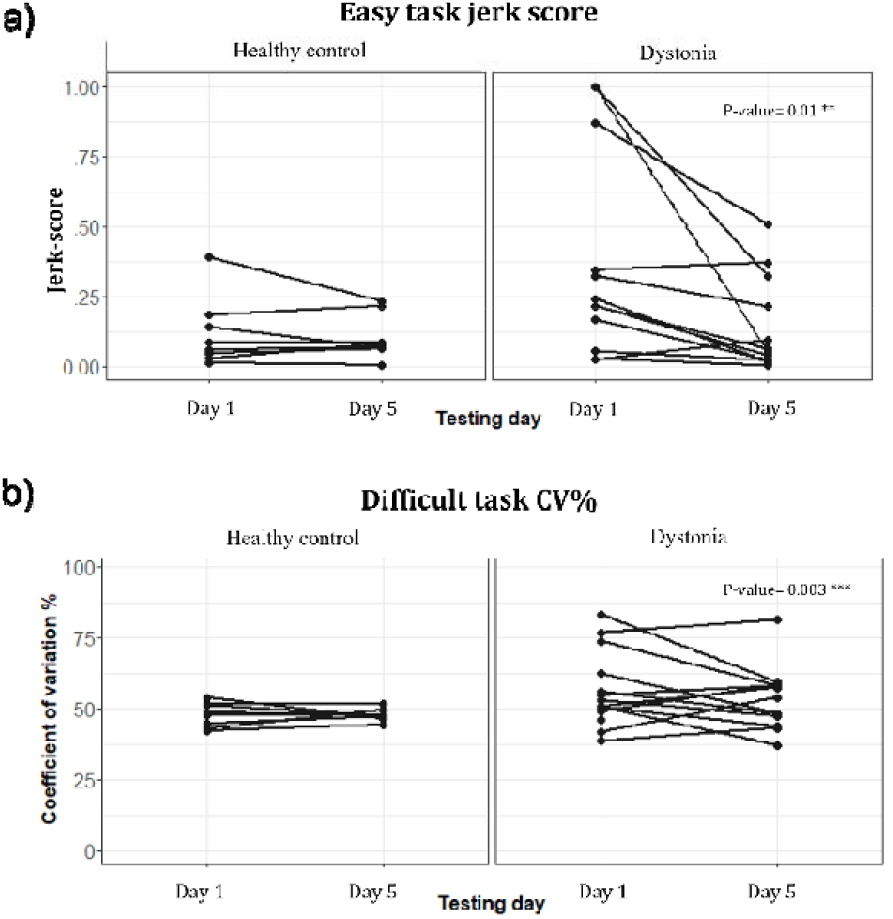
a) Effect of practice on the jerk-score in healthy and dystonia group. b) Effect of practice on the Coefficient of Variation in healthy and dystonia group.

Similar analysis was done on the coefficient of variation of movement in both groups. The models explained 30% and 51% or variance of CV in healthy control and the dystonia group, respectively. The pairwise comparison of mean in group analysis showed there is only a significant change of CV in difficult task (highest ID) in children with dystonia (p-value = .003), however this slope is only significant (p-value < 0.05) for 6 participants out of 13 as shown in Figure 10b (Day 1 mean = 56.5, Day 5 mean = 53).

## Discussion

In this study, we found that children with secondary dystonia and healthy children were able to improve their performance with practice on a trajectory task constrained only by the risk of dropping the marble. This constraint imposes a limitation on the maximum variability during the task performance, without specifying a particular desired trajectory or endpoint accuracy. It may therefore more closely reflect some aspects of normal movement behavior when tracking or tracing is not the purpose. For example, it may represent some of the challenges faced by children as they attempt to feed themselves from a spoon.

Improvement in performance is reflected in improvement in the speed accuracy tradeoff, as measured by increased speed for each level of difficulty, as well as IP. In order to increase speed without dropping the marble, it is necessary to reduce the movement variability within each trajectory, or to decrease the maximum acceleration and jerk of the spoon that would lead to the marble exceeding the bounds of the spoon edge.

We found that smoothness (evaluated by jerk-score) and coefficient of variation improved significantly in children with dystonia but not in healthy children. Although we would have expected a decrease in healthy children, in fact some of their improvement in performance may have occurred due to a decrease in their initial large e_1_ safety margin. In other words, healthy children tended to maintain the marble closer to the center of the spoon, thus perhaps not achieving the maximum speed that could be achieved had they allowed the marble to approach the edge of the spoon. With practice, greater confidence in performance may have allowed them to reduce the safety margin and allow higher levels of variability without exceeding the bounds of the spoon. In contrast, children with dystonia had much less of a safety margin at baseline, perhaps due to their higher intrinsic variability. The only way that children with dystonia could thus improve performance would be to improve the smoothness itself, because the safety margin could not be safely reduced without dropping the marble.

In conclusion, we have shown that both healthy children and children with secondary dystonia improve skill as measured by the speed-accuracy tradeoff on a trajectory task where variability is constrained by the physics of the task rather than adherence to a target trajectory. While improvement of performance with practice is not surprising in healthy children, it is nevertheless interesting to note that this is reflected in a change in the speed-accuracy tradeoff, indicating that Fitts’ law is not immutable but rather represents the current level of skill and task performance^1,2,7^. Improvement of performance in children with secondary dystonia is interesting because this suggests that the higher level of signal-dependent noise can be controlled through repetition and learned strategies, and this provides an avenue for the quantitative evaluation of rehabilitation strategies in this otherwise highly challenging group^3–5,13^.

The significant increase in the index of performance arises from different approaches in the two different subject groups. Healthy children improved by reducing the safety margin, and perhaps maintaining the same level of signal dependent noise, whereas the children with dystonia maintained the same safety margin but reduced their noise and movement variability. Children with dystonia appropriately adjust their speed to compensate for the level of variability, consistent with prior results^21^. Prior research has shown that the origins of signal dependent noise may be different in these two groups, and perhaps only the noise in the children with dystonia is amenable to reduction with practice^5,21^. Further study of modifications of the speed-accuracy tradeoff in this and other task are warranted in order to evaluate the potential for improvement in skill with practice in children with secondary dystonia.

The limitations of this work include the random choice of task difficulties for each subject, however we confirmed that this variable did not have a significant effect on the final results by comparing the outcome variables with respect to both spoon number (task difficulties; easy, medium, and difficult) and ID. In addition, due to the different capabilities of participants, we observed ceiling effect in the speed of movement in some subjects, however this was only limited to some repetitions and the effect was cancelled out by fitting linear models.

## Data Availability

All data produced in the present study are available upon reasonable request to the authors

## Acknowledgements

We would like to acknowledge Ambra Cesareo, Claudia Casellato, Andrea Galbiati, Shinchi Amano, Jonathan Realmuto, and Ritt Givens for their support to data acquisition and Aprille Tongol and Elena Beretta for their help in patient recruitment. This work was supported by the US National Institute of Health (grant R01HD081346-01A and Subaward USC-POLIMI: 61430868) and by the Italian Ministry of Health (Ricerca Corrente 2015/2022 to E. Biffi).

